# Characterising subgroups of people with severe COVID anxiety by latent profile analysis

**DOI:** 10.1101/2023.04.28.23289248

**Authors:** Jacob D King, Aisling McQuaid, Verity C Leeson, Oluwaseun Tella, Mike J Crawford

## Abstract

**Background:** Severe COVID anxiety describes people whose experiences of the COVID-19 pandemic are overwhelming, and have lead to patterns of behaviours that add little protective benefit but are at the expense of other priorities in life. It appears to be a complex social and psychological phenomenon, influenced by demographic and social factors. Identifying subgroups of people with severe COVID anxiety would better place clinicians to assess and support this distress where indicated.

**Methods:** Measurement tools assessing depression, generalised and health anxiety, obsessive-compulsive symptoms, personality difficulty and alcohol use from 284 people living in United Kingdom with severe COVID anxiety were explored with latent profile analysis. Further analyses examined the associations of identified clusters with demographic and social factors and daily functioning, quality of life and protective behaviours.

**Results:** A model with 4 classes provided the best fit. Distinct patterns of psychopathology emerged which were variably associated with demographic factors and COVID behaviours.

**Limitations:** Given the complex aetiology of COVID anxiety a number of factors which might better cluster subgroups are likely to have gone uncollected. Moreover, using data collected at a single time-point limits these results’ ability to conclude whether observed relationships were the product of the pandemic or longstanding.

**Conclusions:** People living with severe COVID anxiety are a heterogenous group. This analysis adds to evidence that certain health behaviours and demographic factors are inextricably linked to poor mental health in people with COVID anxiety, and that targeting health behaviours with specific intervention might be beneficial.

## Introduction

Anxious thoughts and feelings about the COVID-19 pandemic are common, and for most people encourage the use of appropriate protective behaviours like hand washing, isolating if symptomatic and testing (Pierce et al., 2021). However for some people fears of the virus are persistent and overwhelming, are associated with unpleasant somatic symptoms sometimes misattributed to COVID-19 itself, and behaviours intended to be protective but are in far excess of any public health guidelines and stop people from getting on in their life (Arora, Jha, Alat, & Das, 2020). Figures from a nationally representative sample of the German population suggest around 70% of people do not report any physiological symptoms of anxiety when exposed to a reminder about the virus, however for 5% these experiences are severe, and associated with significant day-to-day dysfunction (Hajek & König, 2022). In North America up to 16% of the surveyed population experienced significant anxious responses in relation to fears of COVID-19 and the impacts to society, which authors highlight might benefit from mental health support (Taylor, Landry, Paluszek, Fergus, et al., 2020). However to date very little work has been conducted on the subgroup of people with this clinically important and persistent level of COVID anxiety, nor the role that mental health services might play in providing support.

Anxious responses in relation to a COVID-19 stimulus appear to be a heterogenous experience, and a myriad of assessment scales have been developed to assess these thoughts, feelings and behaviours (Ahorsu et al., 2022; Lee, 2020; Ana V. Nikčević & Spada, 2020). Studies to date recognise the role of fear of contagion for oneself and loved ones, and worries about the consequences of the pandemic including socioeconomic impacts and an overwhelmed healthcare system (Asmundson et al., 2020; Taylor, Landry, Paluszek, & Asmundson, 2020). Multiple formulations are described and the contribution of generalised, social, and health anxiety, traumatic stress, personality, and obsessive-compulsive symptomatology is recognised (A. V. Nikčević, Marino, Kolubinski, Leach, & Spada, 2021; Taylor, Fong, & Asmundson, 2021). Other researchers have suggested that an enhanced risk of hospitalisation or death, or living with someone at increased risk from COVID-19, perhaps due to age, ethnicity or an at-risk health condition leads to a greater burden of anxious responses, appropriately in the context of the significant morbidity and mortality risk posed by the virus. The loss of loved-ones, personal experiences of ill health due to COVID-19 and the decline of social networks are also all likely to contribute to enhancing anxious feelings, and disrupt established coping mechanisms (Lokman & Bockting, 2022).

Public health bodies globally have recommended adopting protective behaviours to reduce transmission of SARS-Co-V2. However government guidelines in the UK in some cases specified that excessive use of these behaviours like washing hands constantly and bleaching or quarantining items coming in to the home, were not necessary (UK Health Security Agency, 2020, 2021), yet these excessive behaviours were common among people with severe COVID anxiety (King et al., 2023). Bish and colleagues, reviewing protective behaviours used in pre-COVID viral pandemics, identified several demographic and psychological factors associated with these behaviours, namely a heightened perception of susceptibility to the virus, being older and female were also predictive of employing protective behaviours (Bish & Michie, 2010). There has now been a huge amount written about the COVID-19 pandemic, and there are strong associations between fear of COVID-19 and poor mental health (Alimoradi, Ohayon, Griffiths, Lin, & Pakpour, 2022) along with evidence that individual level psychological factors might predict dysfunctional COVID-19 fear better than macro-level environmental factors (Eder et al., 2021). However to date, we are not aware of any study among individuals with severe COVID anxiety that has explored whether there are meaningful clinical subgroups, and associations with particular demographic and clinical factors. A better understanding of subgroups within this cohort is likely to help advance understanding of anxious responses to pandemics including protective behaviours, inform causative networks of these experiences and behaviours, and better inform effective support to those with severe anxiety in the context of the all too real risks COVID-19 poses to health.

## Methods

This article reports a secondary analysis of data collected at baseline from Imperial College London’s COVID Anxiety Project – a protocol for which is published previously (Crawford et al., 2022). Initially over 1,000 individuals responded to recruitment adverts placed on social media platforms and contact from London based primary care clinics, of which 306 scored 9 or more on the COVID Anxiety Scale (CAS) (Lee, 2020) indicating severe COVID anxiety, consented, and met other eligibility criteria including living in UK, being aged over 18 and not having a history of psychosis.

Ethical approval for collection of data was granted by the Leicester Central Research Ethics Committee and Health Regulation Authority in 2020, reference number 20/EM/023, and was conducted in accordance with Declaration of Helsinki 1964. All data management and analyses were performed using STATA IC 16.1. All statistical significance testing was 2-sided.

### Model development

We worked with a medical model of severe COVID anxiety, presupposing that symptoms of mental illness (depression, generalised anxiety for example) with their complex biological, social and psychological underpinnings, formed the initial impetus from which severe COVID anxiety symptoms, and subsequently poor quality of life, difficulties with daily living, and health behaviours resulted. We planned for mental health conditions which are suspected to contribute to the psychopathology of severe COVID anxiety to be included as indicator variables in a latent profile analysis (LPA) to explore if there were meaningful subgroups of individuals living with severe COVID anxiety. LPA is a finite mixture model-based clustering approach which operates on the probability of datapoints relating to each other through a hidden ‘latent’ class. It is a “person-centred” approach, grouping individuals based on similarities rather than other “variable-centred” approaches which could be less clinically useful, contracting cut-offs with an abstracted real world meaning. Indicator variables were selected *a priori* based on literature review and clinical experience. We intended to explore whether there were any differential associations between clusters and demographic covariates and clinical outcomes. Respectively these included factors contributing to health risk from COVID-19, age, sex, ethnicity and health conditions which increased a person’s risk to life from COVID-19, and quality of life, social and occupational functioning, and COVID-19 specific protective behaviours.

### Measures

Demographic factors were collected including age, sex, ethnicity, living arrangements (alone or with others, and living with someone thought to be at risk from COVID-19 or not), employment status, previous COVID-19 infection, whether a loved one had been hospitalised by COVID-19, along with a battery of psychopathological self-complete screening tools to measure depression, generalised anxiety, health anxiety, obsessive-compulsive disorder, personality difficulty and alcohol use. These were measured respectively with the Patient Health Questionnaire – 9 items (PHQ-9), Generalised Anxiety Disorder scale – 7 items (GAD-7), the 14 health anxiety items of the Short form Health Anxiety Inventory (sHAI), Obsessive-Compulsive Inventory – Revised (OCI-R), the Standardised Assessment of Personality Abbreviated Scale (SAPAS), and the Alcohol Use Disorders Identification Test for Consumption (AUDIT-C). The presence or absence of an at-risk health condition was based on participant self-reports of their medical conditions. These responses were coded using the MedDRA system (MedDRA, 2022) by a physician, and at-risk conditions were identified by those highlighted in the QCOVID study (QCovid Risk Assessment, 2022). Ethnicity was dichotomised by whether the respondent reported belonged to a UK Census ethnic group at an increased risk from COVID-19: Black (Black Caribbean/African), South Asian (Indian, Pakistani, Bangladeshi) and ‘Other’ groups, compared to White British and Irish, White Other and Chinese background.

Outcomes of interest included social and occupational functioning as measured by the Work and Social Score (WSAS), health-related quality of life as measured by the EuroQuol 5 dimensions 3-item scale (EQ-5D-3L), and COVID specific health related behaviours. These COVID health behaviours included frequency of hand washing and clothes washing, washing disinfecting or quarantining items brought into the home, the frequency of leaving one’s home, and consuming COVID related news, each mounted on 4 or 5 point Likert scales. For each behaviour one of the options represented an extreme variant of the behaviour, which in some cases had been explicitly confirmed by public health bodies to be in excess of guidance. Likert scales were dichotomised: excessive variant of the behaviour versus not excessive variant. EQ-5D-3L scores were converted to a continuous variable using United Kingdom specific population standardised scores with the Time-Trade Off approach (Devlin, Parkin, & Janssen, 2020). A full description of included variables along with cut-off scores and explanations of variable generation have been described in previous articles from the COVID Anxiety Project (King et al., 2023).

In order to facilitate clear interpretation continuous scores from psychopathology screening tools were standardised by z-score. This was also favourable to dichotomising the presence or absence of the disorder with screening diagnostic cut-off values due to very high rates of generalised anxiety (91.5%) and depression (85.5%): those without these disorders made up a very small contingent of the sample with around 25 and 40 participants respectively. Outliers of categorical variables were assessed graphically, aside the AUDITC no scales featured outliers, and outliers were retained in AUDITC due to the heavy skew related to a high percentage of the cohort not reporting drinking any alcohol. The assumption of indicator independence was confirmed with a series of pairwise Spearman rank correlation tests, where each was weakly or not at all correlated with the other (r<0.5) aside an expected moderate correlation between GAD-7 and PHQ-9 of 0.59.

### Participants

Analysis was restricted to 284 participants providing complete data. Participants were required to complete all items of individual screening tools before progressing through the interview, as such there are no within-tool missing data. The majority of the sample were female (231, 81.3%) and White British or Irish (70.8%): median age was 41.3, and 14.8% of the sample were aged 60 or older. 52 individuals were of an at-risk ethnic group (17.2%) and 45 (15.9%) lived alone. Health conditions which put the individual at increased risk of hospitalisation from COVID were reported by 25.7%, and at the time of baseline reporting 46 individuals (16.2%) with severe COVID anxiety reported previous COVID infection. A full description of baseline data is reported elsewhere (King et al., 2023).

## Results

We used a standard 3-step approach as outlined by Vermunt (2017). The model was developed using STATA’s *gsem* command. We drew 50 random starts in attempt to avoid local solutions, and ran with 100 iterations. We ran model variants permitting the error terms’ variance to be unequal across classes, and error terms to be correlated across classes.

### Model selection

We first fit 1 to 6 latent profile models incrementally. The most appropriate number of classes was selected based on theoretical plausibility and clinical interpretability, class size (not less than 10% of the sample), the lowest Aikaike (AIC) and Bayesian (BIC) information criterion, and the average posterior class probabilities (AvPP), where greater than 0.7 represents an acceptable level of accuracy for classification of individuals within each class. The entropy of models, which expresses how distinct clusters are from each other was also calculated - an entropy >0.8 is assessed as ‘good’ separation (Weller, Bowen, & Faubert, 2020). Table 1 outlines the fit statistics of these models. The model with 4 clusters was selected based on considering the models with lowest information criteria, highest posterior predicted probability, and a preference for clinical interpretability and parsimony. While the model with 3 clusters provided slightly more favourably low information criteria, the 4 cluster model provided notably more meaningful insights and was therefore preferred (Berlin, Williams, & Parra, 2013; Spurk, Hirschi, Wang, Valero, & Kauffeld, 2020).

Secondly, we allocated individual respondents to clusters by their posterior class membership probabilities. Before lastly using these allocated cluster variables in further analyses: multinomial logistic regression was used to identify differences in demographic and lifestyle factors between clusters, and one-way ANOVA was used to identify differences between classes and distal outcomes, which are reported in Tables 2 and 3 respectively. Full code and dataset will be made available from the COVID Anxiety Project in public repositories on completion.

### Clusters

The first group representing 31.7% (n=90) of the sample scored significantly lower on all screening measures than the whole cohort mean, with group means of health anxiety, OCD and personality difficulty hovering around diagnostic thresholds for the scales used. Moreover this cluster with a more “simple” severe COVID anxiety reported the highest average quality of life of the sample approaching UK national average levels (Janssen et al., 2019), a group average ‘moderate’ level of social and occupational dysfunction and below cohort average rates of each of the measured excessive protective behaviours.

The second and largest cluster of people living with severe COVID anxiety (37.7%, n=107) did not differ in their profiles on scores for depression, generalised anxiety, health anxiety, obsessive compulsive symptoms and personality disturbance from the cohort average, however no member of this cluster reported alcohol consumption, returning cluster mean AUDIT-C score of 0. Importantly, this group were more likely to report a diagnosis of a condition which increased their risk of hospitalisation or death from COVID-19 (p=0.043), which was present in 34.5% of this cluster compared with an average of 25.7% in the rest of the sample. They were the oldest median aged group (aged 44), and had the lowest percentage of men of any cluster (12%). This group were the most likely to report never leaving their home (16.8%) since the pandemic started. Half of this group also continued to buy their shopping entirely online, and 40% continued to clean all groceries entering the home, both notably higher than other clusters.

The third grouping (11.3%, n=32) clustered with greater than average rates of obsessive-compulsive symptoms and personality difficulty, but cohort average severity of depressive and generalised anxiety symptoms, and hence labelled ‘anakastic’. This group were the youngest median aged cluster (36 years old), and men and people from an at-risk ethnic background were represented at twice the rates of the whole cohort.

The final group (19.4%, n=55) demonstrated significantly higher scores on all psychopathological assessment scales apart from alcohol use. This cluster with the most “complex” severe COVID anxiety were more likely to report living with someone thought to be vulnerable to COVID-19 (49.1%, p=0.016), as well as the group most likely to be report living alone (23.6%, p=0.007), and least likely to be in employment. These individuals who clustered with the highest scores on psychopathological outcome measures also experienced the worst average rates of social and occupational dysfunction, a very poor quality of life, and were significantly more likely to report watching COVID-19 news media ‘constantly’ and washing their hands ‘constantly’.

## Discussion

This latent profile analysis of a sample of 284 UK adults living with severe COVID anxiety has shown distinct and clinically meaningful groupings. While high rates of co-occurring psychopathology were apparent in this population as a whole, LPA identified specific latent patterns. A contingent of a relatively ‘simple’ severe COVID anxiety emerged at one end, along with a more severe complex grouping with significant cooccurring psychopathology at the other. Two further groups emerged, one with a distinctive ‘anakastic’ profile characterised by a high burden of obsessive-compulsive and personality pathology, as well as a group abstinent from alcohol with the highest average age and higher than expected rates health conditions which increased risk to life from COVID-19.

There are several key findings from this analysis which add to current understandings, providing evidence to refine models of COVID anxiety.

First, it appears that demographic and lifestyle factors, and excessive health behaviours are associated differentially by psychopathological clusters, raising questions about the way that COVID anxiety, symptoms of other mental state disorders, demographic factors and health behaviours interact with each other. For example research very early in the pandemic identified that health anxiety, frequent media consumption and concerns with the health of loved-ones were the greatest predictors of greater COVID-19 related fear at that initial point (Mertens, Gerritsen, Duijndam, Salemink, & Engelhard, 2020). The complex severe COVID anxiety cluster identified in this analysis demonstrated co-associations between constantly consuming COVID-19 media and living with someone thought to be vulnerable to COVID-19. So while these factors appear to be predictors of greater levels of COVID anxiety, they also cluster together in people with severe COVID anxiety and universally poorer mental health, social functioning and quality of life outcomes.

In conjunction, previous regression analyses of this dataset established factors most predictive of certain health behaviours - for example generalised anxiety symptoms and living with someone thought to be vulnerable to COVID-19 predicted consuming media, which has clustered together in this present analysis (King et al., 2023). However increasing OCD symptomatology predicted more severe hand washing behaviours, yet between the two clusters with higher than expected OCD symptoms - the anakastic and complex severe COVID anxiety groups - the complex psychopathology group features twice the rates of constantly washing hands, perhaps signalling that greater OCD symptoms alone might not be sufficient for employing this behaviour dysfunctionally, and that other social or demographic factors come in to play.

The abstinent group clustered with the oldest average age, highest proportion of female members and a greater prevalence of at-risk conditions: these demographic covariates have been variably associated with the employment of protective behaviours to an excessive degree in other datasets (Cao, Siu, Shek, & Shum, 2022; Caycho-Rodríguez et al., 2022; Miola et al., 2023; Ye et al., 2020). This cluster also demonstrated the highest rates of certain health behaviours, with more than half shopping exclusively online due to COVID fears, and the greatest rates of never leaving home and disinfecting all items coming in to their homes. With cohort average rates of psychopathology it appears that the three demographic factors associated with an abstinent psychopathology profile, in turn associate with these particular health behaviours. The association of at-risk characteristics and certain health behaviours have been played out in research extensively during this pandemic (Bish & Michie, 2010) and this clustering analysis appears to replicate these findings among individuals with severe COVID anxiety abstinent of alcohol.

This analysis also highlights the importance of social environment for people with COVID anxiety. The complex psychopathology group were both the most likely to report living with someone thought to be vulnerable to COVID-19, but also the group most likely to report living alone. We find the evidence pointing to the ‘emotional contagion’ of fear of the pandemic, that is, that living with someone vulnerable to COVID-19 or themselves experiencing COVID anxiety might perpetuate one’s own fears, to be convincing in explaining this former association (Kojan, Burbach, Ziefle, & Calero Valdez, 2022; Wheaton, Prikhidko, & Messner, 2020) and cite other evidence linking social isolation to poorer mental health outcomes during the pandemic as supporting the latter (Bzdok & Dunbar, 2022). We therefore might argue that the complex psychopathology group, may have clustered as such due to the presence of social environmental factors (living alone, or with someone thought to be vulnerable to COVID), and the influence of ‘constant’ hand-washing and media consuming behaviours, all together creating a milieu contributing to poor outcomes.

Finally, relating to the role of health anxiety in severe COVID anxiety, a number of research groups appear to have arrived at similar conclusions, that health anxiety processes are a core facet in the psychopathology of COVID anxiety (Alimoradi et al., 2022; Asmundson & Taylor, 2020; Crawford et al., 2022; A. V. Nikčević et al., 2021) relating instrumentally to factors including perceived risk and threat-selective attention. In the present analysis health anxiety scores did not appear to discriminate between clusters, aside from the simple cluster who varied somewhat, while the others maintained near cohort average severities of health anxiety. This finding is consistent with health anxiety psychopathology featuring as a common facet to otherwise different underlying psychopathological profiles of people living with severe COVID anxiety.

Together these findings are likely to influence proposed models of COVID anxiety, associated mental health and protective behaviours. While factors which cluster together do not indicate causal relationships, given that psychopathological clusters associate with demographic and health behaviours in a specific pattern, there are likely complex interactions at play. Case in point, consuming COVID-19 media and living alone or with someone thought to be vulnerable to COVID-19 could operate bi-directionally with COVID anxiety, reinforcing each other, and thus these analysis add further support to the hypothesis that targeting health behaviours especially the high consumption of COVID-19 media could be an effective intervention for reducing poor mental health among those with COVID anxiety (Elhai, Yang, McKay, & Asmundson, 2020; Tyrer, 2020; Wu, Nazari, & Griffiths, 2021).

While in some longitudinal analysis “fear of coronavirus” appeared to reduce for some of the general population over time, other stressors like social isolation, job insecurity, and worries about the future continued after levels of the circulating virus had declined (Leung & Khalvati, 2022). The COVID Anxiety Project plans to collect follow-up data on people in this cohort 18 months after their recruitment, which will be able explore the longitudinal outcomes of each identified cluster and factors associated with remission of symptoms.

### Strength and limitations

A key benefit of this study is its focus on people with severe COVID anxiety, permitting an analysis of factors which associate with poor mental health, social functioning and quality of life outcomes in this clinically important group. One previous categorisation approach measuring mostly mild COVID anxiety as a continuous trait had fallen to the so-called ‘salsa effect’, a situation in which clusters organise themselves into a ‘mild’, ‘moderate’, ‘severe’ pattern along a unidimensional axis rather than identifying subgroups per se (Mani et al., 2022). This present analysis was able to identify more clinically meaningful groupings.

The study has a number of limitations due to the cross-sectional design. Many mental health outcomes deteriorated among the general population during the COVID-19 pandemic, and while some evidence suggests that people already living with poor mental health were more likely to experience severe COVID anxiety (Asmundson et al., 2020), we cannot be sure from this present analysis whether these clusters, for example the anakastic group, existed in their symptom profiles before COVID-19, or whether these factors particularly OCD symptoms and alcohol use, emerged as a response to COVID-19 threat. Moreover this study was limited to adults living in the UK, and sampled only a period during the second and third waves of the pandemic. The sample was self-selecting and while demographically diverse was not a selected representatively or randomly. While this approach allowed us to select for people with persistent COVID anxiety, dynamic temporal and regional factors which may have had an impact on several assessment measures ad behaviours could not be adjusted for. Other factors which may have mediated the relationships between severe COVID anxiety, mental health symptoms and behaviours, particularly personality traits, were also not collected (A. V. Nikčević et al., 2021).

## Conclusions

People with severe COVID anxiety are a heterogenous group, represented by varying patterns and levels of severity of psychopathology, health behaviours and demographic factors. These data support a model of COVID anxiety in which health anxiety is a core facet, and where specific psychopathological profiles are associated variably with demographic factors, protective behaviours, poor quality of life and social and occupational dysfunction. Future work which looks at the trajectory of these clusters, along with examining any differential benefits of interventions on these clusters would be of benefit to support people still experiencing severe COVID anxiety, and to predict responses in any future pandemic-like events.

***

## Data Availability

Data relating to the present analysis are available from the corresponding author on reasonable request. In due course the full COVID Anxiety Project dataset will be made publicly assessable.

## Ethics approval and consent to participate

The COVID Anxiety Project received approval from the Leicester Central Research Ethics Committee and Health Regulation Authority in 2020, reference number 20/EM/023. Research was conducted in line with the Declaration of Helsinki 1964, and responses were managed following General Data Protection Regulations. All individuals were required to provide informed consent, by independently reading a participant information sheet before signing an online consent form to participate. Participants received a copy of their consent form.

## Authors’ contributions

All authors meet ICJME criteria for authorship. JDK drafted initial versions of this manuscript. MJC and VCL were involved in the design of the COVID Anxiety Project. AM and JDK collected data and VCL and OT coordinated the study. JDK led on data analysis, and had full access to raw data. All authors reviewed and approved a final version of this manuscript.

## Acknowledgements

We wish to thank members of the lived experience reference group including Manisha Ahya, Charlotte Green, Sandra Jayacodi, Niruben Patel and Vikas Sharma for their contributions to contextualising the results of this study and in the development of novel COVID-19 health behaviour items.

