## Supplementary material for "Characterising subgroups of people with severe COVID anxiety by latent profile analysis": Tables

| **Fit statistic** | **1 Class** | **2 Class** | **3 Class** | **4 Class** | **5 Class** | **6 Class** |
| --- | --- | --- | --- | --- | --- | --- |
| Log likelihood-ratio | -2261.81 | -2219.80 | -1572.60 | -1570.81 | -1655.90 | -2100.47 |
| AIC | 4577.61 | 4549.59 | 3303.21 | 3353.62 | 3557.80 | 4534.93 |
| BIC | 4676.14 | 4750.29 | 3591.48 | 3740.41 | 4006.62 | 5144.31 |
| Entropy | 1.0 | 0.71 | 0.99 | 0.99 | 0.90 | 0.88 |
| Smallest class (%) | 100 | 43.3 | 13.0 | 11.3 | 3.5 | 8.1 |
| AvPP | 1.0 | 0.93 | 0.85 | 0.88 | 0.89 | 0.96 |

Table 1. Information criteria and fit statistics. AIC = Akaike Information Criteria, BIC = Bayesian Information Criteria, AvPP = Average Posterior class Probability.

| N=284 | Whole cohort [mean, s.d./IQR] | Cluster 1  ***‘Simple’ severe COVID Anxiety*** | Cluster 2  ***Abstinent*** | Cluster 3  ***Anakastic*** | Cluster 4  **‘Complex’ severe COVID anxiety** |
| --- | --- | --- | --- | --- | --- |
| Indicators | | z-score  [mean, s.d.] | z-score  [mean, s.d.] | z-score  [mean, s.d.] | z-score  [mean, s.d.] |
| **GAD** | [15.5, 3.96] | –0.46***  (13.5, 3.9) | –0.02  (15.4, 4.0) | 0.14  (16.1, 2.9) | 0.79***  (18.6, 2.1) |
| **PHQ** | [15.7, 5.4] | –0.34**  (13.7, 5.2) | –0.05  (15.4, 5.6) | –0.11  (15.1, 3.5) | 0.77***  (19.8, 3.8) |
| **sHAI** | [23.5, 7.0] | –0.25**  (21.5, 5.9) | 0.15  (24.5, 7.2) | –0.22  (21.6, 5.7) | 0.20  (25.2, 8.0) |
| **OCI** | [30.1, 15.3] | –0.49***  (21.5, 12.0) | –0.13  (28.0, 13.8) | 0.74***  (40.8, 12.0) | 0.80***  (42.5, 13.0) |
| **SAPAS** | [4.2, 1.84] | –0.35**  (3.4, 1.7) | –0.12  (3.9, 1.9) | 0.45**  (5.0, 1.5) | 0.63***  (5.4, 1.4) |
| **AUDIT-C** | [2.7, 2.94] | 0.62***  (4.5, 2.5) | –0.91***  (0, 0) | –0.23***  (2.0, 0.8) | 0.86***  (5.4, 2.7) |
| Covariates | | %/median, (Beta-coefficient) | %/median, (Beta-coefficient, p) | %/median, (Beta-coefficient, p) | %/median, (Beta-coefficient, p) |
| **Age** | 41.2 [28.5 – 53.4] | 42.6  (baseline) | 44.8  (0.004, 0.67) | 36.02  (-0.02, 0.20) | 39.3  (-0.02, 0.63) |
| **Sex (male)** | 18.7% | 17.8%  (baseline) | 12.1%  (-0.48, 0.27) | 37.5%  (0.75, 0.15) | 21.8%  (-0.23, 0.63) |
| **At risk ethnic group** | 17.2% | 12.4%  (baseline) | 14.2%  (0.30, 0.51) | 35.5%  (0.89, 0.12) | 20.8%  (0.36, 0.49) |
| **At risk health condition** | 25.7% | 20.0%  (baseline) | 34.6%  (0.71, 0.043*) | 15.6%  (-0.07, 0.90) | 23.6%  (0.38, 0.39) |
| **Living Alone** | 15.9% | 11.1%  (baseline) | 15.9%  (0.49, 0.29) | 15.6%  (0.41, 0.55) | 23.6%  (1.39, 0.007**) |
| **Employed** | 44.7% | 50.0%  (baseline) | 41.2%  (-0.28, 0.36) | 37.5%  (-0.67, 0.14) | 47.3%  (-0.03, 0.94) |
| **Vulnerable cohabitant** | 39.4% | 32.2%  (baseline) | 37.4%  (0.20, 0.54) | 50.0%  (0.53, 0.25) | 49.1%  (0.92, 0.016*) |
| **Loved-one hospitalised by COVID-19** | 33.5% | 33.3%  (baseline) | 34.6%  (0.16, 0.61) | 31.3%  (-0.29, 0.57) | 32.7%  (-0.001, 0.99) |
| **Had COVID** | 16.2% | 21.1%  (baseline) | 14.0%  (-0.57, 0.15) | 9.4%  (-0.89, 0.19) | 16.4%  (-0.33, 0.48) |

Table 2. Profiles of indicators and covariates by class. *p<0.05, **p<0.01, ***p<0.001

|  | Whole cohort mean (s.d.) | Cluster 1  ***‘Simple’ severe COVID Anxiety*** | Cluster 2  ***Abstinent***  Mean/median (s.d.) | Cluster 3  ***Anakastic***  Mean/median (s.d.) | Cluster 4  **‘Complex’ severe COVID anxiety**  Mean/median (s.d.) | Significance test of between group difference, p= |
| --- | --- | --- | --- | --- | --- | --- |
| Outcomes | | mean/median (s.d.) | mean/median (s.d.) | mean/median (s.d.) | mean/median (s.d.) |  |
| **WSAS** | 21.3 (7.5) | 19.3 (6.6) | 21.3 (7.7) | 22.3 (7.0) | 23.7 (8.2) | 0.007** |
| **EQ-5D-3L index score** | 0.585 (median)  IQR [0.255 – 0.770] | 0.725 (0.434) | 0.516 (0.470) | 0.707 (0.566) | 0.291 (0.596) | <0.001*** |
| Health behaviours | | % of class (sd) | % of class (sd) | % of class (sd) | % of class (sd) |  |
| **Worrying constantly** | 22.2% | 17.8% (0.38) | 26.2% (0.44) | 12.5% (0.34) | 27.3% (0.45) | 0.27 |
| **Washing hands constantly** | 20.4% | 11.1% (0.32) | 23.3% (0.43) | 15.6% (0.37) | 32.7% (0.47) | 0.012* |
| **Washing all items entering the home** | 31.7% | 24.4% (0.43) | 40.1% (0.49) | 25.0% (0.44) | 30.9% (0.47) | 0.093 |
| **Shopping exclusively online due to COVID fears** | 38.0% | 28.9% (0.46) | 50.5% (0.50) | 34.1% (0.48) | 30.9% (0.47) | 0.008** |
| **Never leaving home** | 11.3% | 7.8% (0.27) | 16.8% (0.38) | 6.3% (0.25) | 9.1% (0.29) | 0.002** |
| **Constantly consuming media on COVID** | 11.6% | 7.7% (0.27) | 11.2% (0.32) | 3.1% (0.18) | 23.6% (0.43) | 0.009** |

Table 3. Distal outcome measures by one-way ANOVA testing between clusters. *p<0.05, **p<0.01, ***p<0.001
